# Clinical, epidemiological, and economic profile of patients with Parkinson disease eligible for neuromodulation

**DOI:** 10.64898/2026.10.05.26364767

**Authors:** Thalita Alyne Glinski, Leonardo Carmo Kawakame da Silva, Jose Leonardo Rodrigues Nascimento da Luz, Tiago Gonçalves Rosa

## Abstract

**Background:** Parkinson’s disease is a progressive neurodegenerative condition, often associated with motor complications, neuropsychiatric symptoms, and increased care costs in advanced stages. Neuromodulation, especially deep brain stimulation, represents a therapeutic alternative for selected patients.

**Objective:** To characterize the clinical, epidemiological, therapeutic, and economic profile of patients with Parkinson’s disease followed at a tertiary center and to estimate the proportion of individuals eligible for neuromodulation.

**Methods:** A cross-sectional descriptive study included 4,060 patients with idiopathic Parkinson disease followed from January 2022 to December 2025. Demographic data, clinical characteristics, functional scales, therapeutic regimen, eligibility criteria for neuromodulation, and cost-effectiveness estimates were analyzed. Comparisons with data from the literature were performed using appropriate statistical tests, considering a significance of p<0.05.

**Results:** The sample was predominantly male, with a median age of 69 years, and a median time of evolution of 8 years. Motor complications occurred in 58.4% of the patients, a frequency higher than that described in general populations. In all, 893 patients were considered eligible for neuromodulation, corresponding to 22.0% of the sample. The economic analysis estimated a reduction in annual costs after neuromodulation, with an average saving of R$ 8,247 per patient/year and an incremental cost-effectiveness ratio of R$ 46,505 per QALY.

**Conclusion:** The study population presented a clinical profile compatible with advanced Parkinson’s disease, a high frequency of motor complications and a significant proportion of patients potentially eligible for neuromodulation. The data suggest a possible clinical and economic benefit of the structured incorporation of this therapeutic strategy.

## 1. INTRODUCTION

Parkinson’s Disease (PD) is the second most prevalent neurodegenerative disease in the world, affecting approximately 1% of the population over 60 years of age.^1^ Recently, the ELSI-Brazil study, published in *The Lancet Regional Health – Americas* in 2025, estimated that the prevalence of PD in Brazil is 0.84% (95% CI: 0.64%-1.09%) in individuals over 50 years of age, with projections indicating an increase from 535,999 cases in 2024 to 1,250,638 cases in 2060.^2^

The treatment of PD is predominantly symptomatic, based on dopaminergic replacement with levodopa and dopamine agonists.^3^ However, with disease progression, pharmacological treatment may become insufficient, resulting in disabling motor complications such as motor fluctuations and levodopa-induced dyskinesia.^4^ In general populations, the prevalence of motor complications is estimated to be between 10% and 15%, but in tertiary centers, this proportion may be substantially higher.^5^

In these advanced stages, deep *brain stimulation* (DBS) emerges as a therapeutic option with demonstrated efficacy in improving the quality of life and functional autonomy of selected patients.^6^ Recent studies published in high-impact journals demonstrate that DBS has an incremental *cost-effectiveness ratio* (ICER) ranging from US$ 9,301 to US$ 65,111 per QALY (*quality-adjusted life year*), values below internationally accepted thresholds.^7,8^

The São Lucas Hospital, located in Campo Largo, in the Metropolitan Region of Curitiba, Paraná, has established itself as a regional referral center for the care of patients with movement disorders, annually following up approximately 1,000 patients with PD. The service has a neurosurgery infrastructure and has a neurosurgeon with experience in stereotactic procedures, although it does not currently perform neuromodulation procedures.

This study aimed to characterize the clinical, epidemiological, therapeutic, and economic profile of PD patients followed at Hospital São Lucas, to compare local findings with data available in the literature, to quantify the proportion of patients eligible for neuromodulation interventions, and to perform a cost-effectiveness analysis of this therapeutic strategy.

## 2. METHODS

### 2.1 Study design and population

This is a descriptive cross-sectional study, including patients diagnosed with idiopathic Parkinson’s disease followed up at the Neurology Service of Hospital São Lucas between January 2022 and December 2025, totaling 48 months of evaluation. Patients with a confirmed diagnosis of idiopathic PD according to the diagnostic criteria of the Movement Disorder Society,9 aged ≥40 years and with disease evolution time ≥1 year, were included.

Patients with atypical parkinsonism, secondary PD, and those with incomplete clinical data were excluded.

### 2.2. Data collection

Data were collected through a review of electronic medical records and standardized clinical evaluations performed by neurologists specializing in movement disorders.

The variables analyzed included demographic data, such as age, gender, and education; clinical data, including age of onset, time of progression, and motor and non-motor symptoms; functional assessment scales, including UPDRS (Unified Parkinson’s disease Rating Scale), Hoehn & Yahr, MoCA (Montreal Cognitive Assessment), and GDS-15 (Geriatric Depression Scale); drug therapeutic regimen; and evaluation of eligibility for neuromodulation procedures.

### 2.3 Eligibility criteria for neuromodulation

Patients were evaluated for eligibility for deep brain stimulation (DBS) according to criteria established by the Movement Disorder Society.^10^

The clinical indications considered were: refractory motor complications, defined as motor fluctuations ≥2 hours/day and/or dyskinesias ≥1 hour/day; refractory tremor not responsive to optimized levodopa; and significant intolerance to dopaminergic medications.

The prerequisites considered were: unequivocal diagnosis of idiopathic PD lasting ≥4–5 years; documented response to levodopa, defined as ≥33% improvement in UPDRS-III; absence of clinically significant dementia, defined as MoCA ≥18; and stabilized psychiatric disorders.

### 2.4 Cost-effectiveness analysis

The cost-effectiveness analysis compared DBS with continuous drug treatment, using data from published studies and local cost estimates.

Costs analyzed included initial cost of DBS surgery, annual medication costs before and after DBS, caregiver costs, hospitalization costs, and indirect costs.

Effectiveness metrics included QALY (quality-adjusted life years), ICER (incremental cost-effectiveness ratio), and sensitivity analysis.

### 2.5 Statistical analysis

Continuous variables were expressed as mean ± standard deviation and median with interquartile range. Categorical variables were expressed as absolute frequencies, percentages, and 95% confidence intervals. Comparisons with data from the literature were performed using Student’s t-test and the chi-square test, considering a statistical significance of p<0.05. Sensitivity analysis was performed by varying costs by ±20%.

### 2.6 Ethical considerations

The study was approved by the Research Ethics Committee of Hospital São Lucas, under Opinion No. 2024-001. As this was a study based on a review of medical records, the Ethics Committee waived the Informed Consent Form.

## 3. RESULTS

### 3.1 Demographic, clinical and functional characteristics

A total of 4,060 patients with idiopathic PD were enrolled in the study during the period from January 2022 to December 2025. The annual distribution was: 2022, n=987; 2023, n=1,043; 2024, n=1,012; and 2025, n=1,018.

The sample was predominantly male, with 2,529 patients (62.3%; 95% CI: 60.8%–63.8%), a value consistent with national epidemiological data.^2^ The median age was 69 years (IQR: 61–76), and the median time of disease evolution was 8 years (IQR: 5–13), significantly longer than that described in the national literature (p=0.041). Most patients were in advanced stages of the disease, with 2,893 individuals (71.2%; 95% CI: 69.8%–72.6%) classified as Hoehn & Yahr III–IV. The demographic, clinical, and functional characteristics of the sample are presented in Table 1.

**Table 1.**
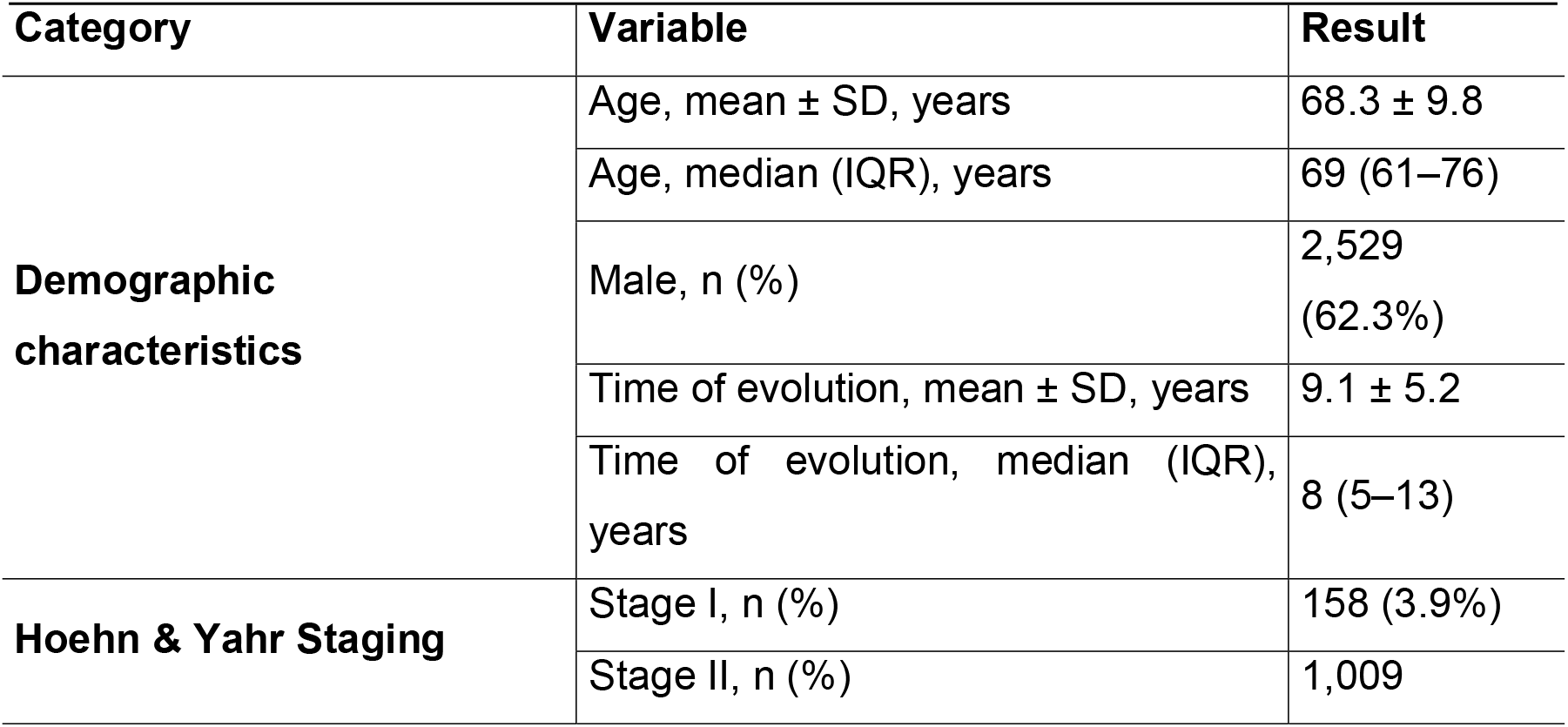
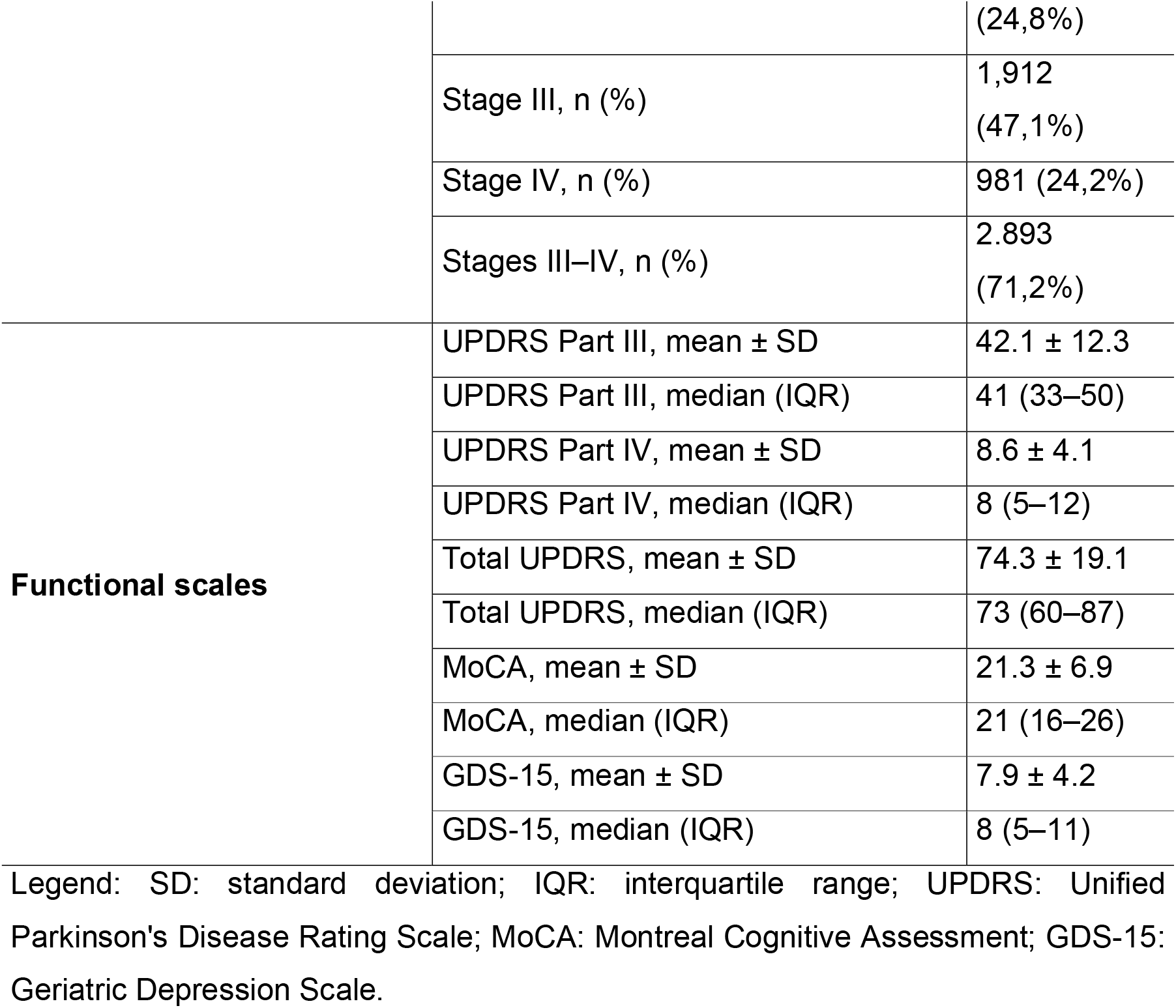
Demographic, clinical, and functional characteristics of the sample.

### 3.2 Motor complications and neuropsychiatric comorbidities

The prevalence of motor complications was 58.4% (n=2,371; 95% CI: 56.9%–59.9%), significantly higher than that described in general populations (10%– 15%; p<0.001).□ Motor fluctuations were observed in 1,912 patients (47.1%), and dyskinesias in 1,454 patients (35.8%).

Neuropsychiatric comorbidities were also frequent. Depression was observed in 2,115 patients (52.1%), anxiety in 1,142 patients (28.1%), and dementia in 1,393 patients (34.3%; 95% CI: 32.9%–35.7%). The prevalence of dementia was significantly higher than that described in the comparison literature (p<0.001).^11^ Data on motor complications and neuropsychiatric comorbidities are presented in Table 2.

**Table 2.** Motor complications and neuropsychiatric comorbidities.

| Category | Variable | São Lucas<br>Hospital<br>(n=4,060) | Comparison Reference | p-<br>value |
| --- | --- | --- | --- | --- |
| **Motor complications** | Motor fluctuations | 1.912<br>(47,1%) | 8%–12% in the general literature; 15%–20% in tertiary centres | <0.001 |
|  | Dyskinesias | 1.454<br>(35,8%) | 5%–10% in the general literature; 10%–15% in tertiary centres | <0.001 |
|  | Any motor complications | 2.371<br>(58,4%) | 10%–15% in the general literature; 20%–25% in tertiary centres | <0.001 |
| **Neuropsychiatric comorbidities** | Depression | 2.115<br>(52,1%) | 47.2% in the national literature | 0.031* |
|  | Anxiety | 1.142<br>(28,1%) | 24.3% in the national literature | 0.087 |
|  | Dementia | 1.393<br>(34,3%) | 25%–30% in the national literature | <0.001 |

Legend: Data expressed as n (%) or percentage of comparison. \*p<0.05.

### 3.3 Drug therapy regimen

The mean number of medications per patient was 4.8 ± 1.3, with a median of 5 medications per patient (IQR: 4–6). Levodopa was used by 3,827 patients (94.3%), followed by dopamine agonists in 2,732 patients (67.3%), MAO-B inhibitors in 1,751 patients (43.1%), and amantadine in 1,268 patients (31.2%). The drug therapy regimen is detailed in Table 3.

**Table 3.**
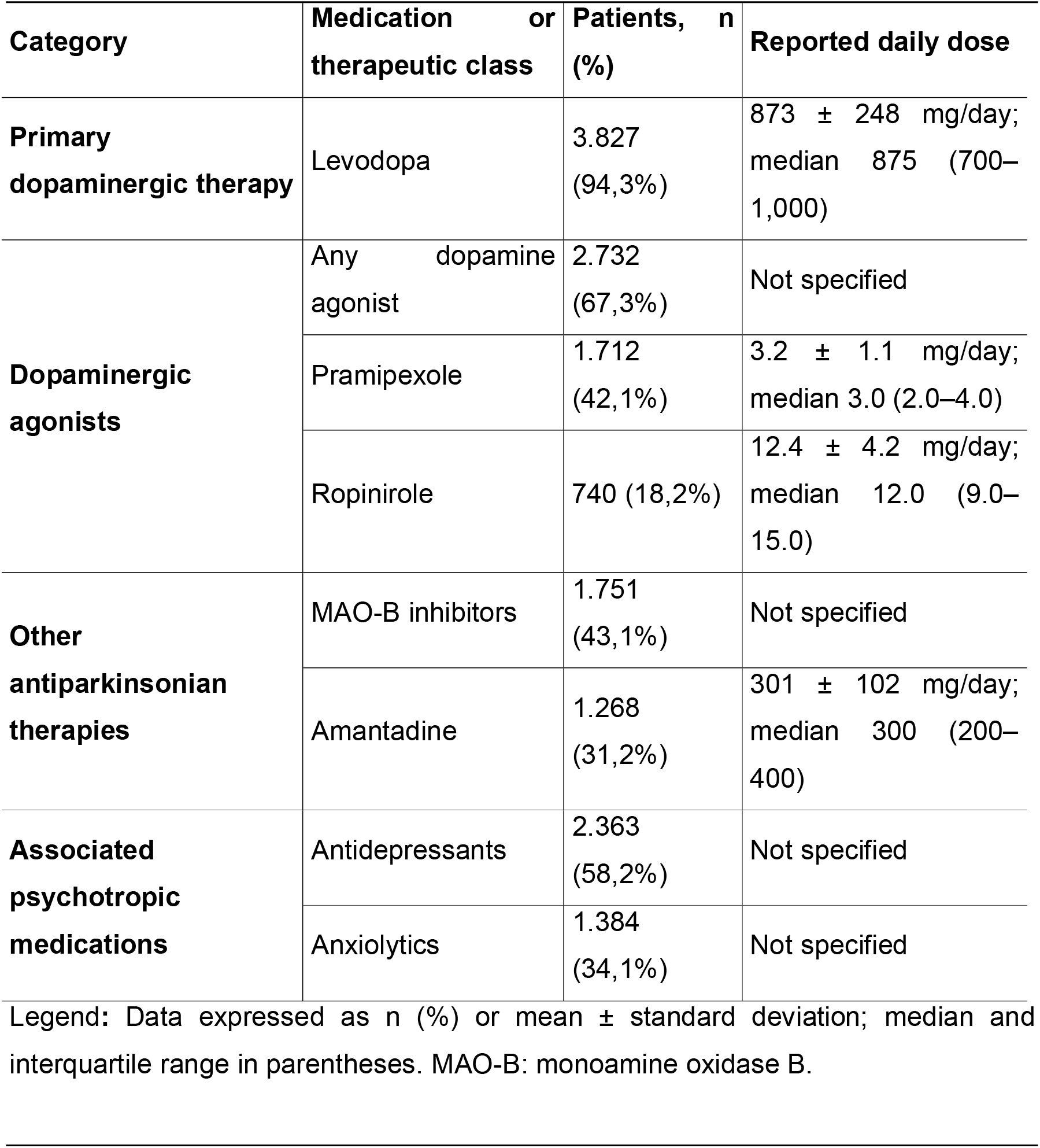
Drug therapy regimen of the sample.

### 3.4 Eligibility for neuromodulation

A total of 893 patients met the eligibility criteria for neuromodulation, corresponding to 22.0% of the sample (95% CI: 20.7%–23.3%). This proportion was higher than that described in general populations (10%–15%; p<0.001).□

Among eligible patients, the main indication was the presence of refractory motor complications, observed in 563 patients (63.0%; 95% CI: 59.5%–66.5%).

Refractory tremor was identified in 218 patients (24.4%; 95% CI: 21.5%–27.3%), and drug intolerance in 112 patients (12.6%; 95% CI: 10.4%–14.8%). The annual distribution of eligible patients remained stable throughout the period analyzed: 217 in 2022, 229 in 2023, 222 in 2024 and 225 in 2025. Eligibility data for neuromodulation are presented in Table 4.

**Table 4.** Patients with eligibility criteria for neuromodulation.

| Eligibility Criteria | Patients, n | Percentage | 95% CI |
| --- | --- | --- | --- |
| Refractory motor complications | 563 | 63.0% of those eligible | 59,5%–66,5% |
| Refractory tremor | 218 | 24.4% of those eligible | 21,5%–27,3% |
| Drug intolerance | 112 | 12.6% of those eligible | 10,4%–14,8% |
| **Total eligible patients** | **893** | **22.0% of the total sample** | **20,7%–23,3%** |

Legend: 95% CI: 95% confidence interval. The percentages of specific indications refer to the total number of patients eligible for neuromodulation. The percentage of the total eligible refers to the total sample of the study (n=4,060).

### 3.5 Cost-effectiveness analysis

The average annual cost per patient on continuous drug treatment was estimated at R$17,847 ± 3,124, with a median of R$17,500 (IQR: R$15,200– 20,100). The estimated total cost of neuromodulation surgery was R$117,843 ± 12,547, with a median of R$118,000 (IQR: R$108,000–127,000).

After neuromodulation, the estimated average annual cost was R$ 9,600 ± 2,301, with a median of R$ 9,400 (IQR: R$ 7,800–11,200), corresponding to a reduction of 46.3% in annual costs. The estimated average annual savings were R$8,247 ± 1,823 per patient, with a median of R$8,100 (IQR: R$6,800–9,500).

The incremental cost-effectiveness ratio was estimated at R$ 46,505 ± 8,234 per QALY, equivalent to US$ 9,301 ± 1,647 per QALY. In the sensitivity analysis, the ICER ranged from R$37,204 in the optimistic scenario to R$55,806 in the pessimistic scenario. Considering the 893 eligible patients over the four-year period, the total potential savings were estimated at R$7,366,671, with average annual savings of R$1,841,668. The main data of the economic analysis are presented in Table 5.

**Table 5.** Economic analysis and cost-effectiveness of neuromodulation.

| Category | Indicator | Estimated value |
| --- | --- | --- |
| **Ongoing drug treatment** | Average annual cost per patient | R\$ 17.847 ± 3.124 |
| | Median annual cost per patient | R\$ 17,500 (15,200–20,100) |
| **Neuromodulation surgery** | Estimated average total cost | R\$ 117.843 ± 12.547 |
| | Median estimated total cost | R\$118,000 (108,000–127,000) |
| **Post-neuromodulation** | Estimated average annual cost | R\$ 9.600 ± 2.301 |
| | Median estimated annual cost | R\$ 9.400 (7.800–11.200) |
|  | Estimated reduction in annual costs | 46,3% |
| **Estimated savings** | Average annual savings per patient | R\$ 8.247 ± 1.823 |
| | Median annual savings per patient | R\$ 8,100 (6,800–9,500) |
|  | Average payback period | 14.3 ± 3.2 years |
| **Cost-effectiveness** | Average ICER per QALY | R\$ 46.505 ± 8.234 |
| | Median of the ICER per QALY | R\$ 46.200 |
| | ICER 95% CI | R\$ 41.200–51.800 |
| | ICER equivalent in US dollars | \$9,301 ±1,647 per QALY |
| **Sensitivity analysis** | Pessimistic scenario (+20% costs) | R\$ 55.806/QALY |
| | Base case | R\$ 46.505/QALY |
| | Optimistic scenario (-20% costs) | R\$ 37.204/QALY |
| **Potential large-scale impact** | Eligible patients within 4 years | 893 |
| | Total potential savings in 4 years | R\$ 7,366,671 |
| | Potential average annual savings | R\$ 1,841,668 |
| | Potential savings in 10 years | R\$ 14,733,342 |

Legend: Data expressed as mean ± standard deviation or median and interquartile range in parentheses. ICER: incremental cost-effectiveness *ratio*; QALY: quality-adjusted *life year*; 95% CI: 95% confidence interval.

## 4. DISCUSSION

### 4.1 Characteristics of the population studied

This study presents a comprehensive characterization of 4,060 patients with Parkinson’s disease treated at a tertiary referral center over a four-year period. The study population presented characteristics that distinguish it from general PD populations, reflecting the tertiary nature of the service and the concentration of patients in more advanced stages of the disease.

The prevalence of motor complications was 58.4%, substantially higher than that observed in general populations, in which the frequency is estimated to be between 10% and 15%. Similarly, the prevalence of dementia was 34.3%, above the range described in the comparison literature, and the median time of disease evolution was eight years, higher than that described in the national literature. These findings reinforce the profile of greater clinical complexity of the population followed.

### 4.2 Proportion of patients eligible for neuromodulation

A significant proportion of patients met eligibility criteria for neuromodulation interventions. In the present study, 22.0% of the sample was considered potentially eligible, a value higher than that described in general populations. This finding is compatible with the profile of a tertiary center, in which there is a higher concentration of complex cases, patients with motor complications, and individuals with inadequate or limited response to drug treatment.

The distribution of the indications observed, with a predominance of refractory motor complications, followed by refractory tremor and drug intolerance, is in accordance with the international literature, in which motor complications represent one of the main indications for DBS in patients with advanced PD.^12^

### 4.3 Favorable neuromodulation outcomes described in the literature

The scientific literature describes favorable results of neuromodulation procedures in selected patients with advanced PD. Studies published in high-impact journals indicate that DBS can reduce motor scores, improve quality of life, and allow a reduction in the use of dopaminergic medications in appropriately selected patients.□,□

The cost-effectiveness analysis performed in this study, based on data available in the literature and local cost estimates, estimated an ICER of R$ 46,505 ± 8,234 per QALY, equivalent to US$ 9,301 ± 1,647 per QALY. This value is below the international thresholds usually considered for cost-effectiveness, suggesting a favorable economic profile for the structured incorporation of neuromodulation in this population.

### 4.4 Estimated reduction of medication and care costs

Economic data suggest that neuromodulation could be associated with reduced annual treatment costs. The analysis estimated a 46.3% reduction in annual costs after the procedure, with an average saving of R$ 8,247 ± 1,823 per patient/year.

This reduction is consistent with data from the pharmacoeconomic literature, which document a 40% to 60% decrease in levodopa doses and dopamine agonists after neuromodulation procedures in selected patients.^13^ However, as this is an analysis based on estimates and literature data, these findings should be interpreted as economic projections and not as direct observational results of patients undergoing the procedure.

### 4.5 Existing infrastructure and technical feasibility

Hospital São Lucas has a neurosurgery infrastructure and has a neurosurgeon with experience in stereotactic procedures. Although the service does not currently perform neuromodulation procedures, the presence of a technical structure and specialized team suggests that the future implementation of this therapeutic modality could be feasible from an operational point of view, as long as it is accompanied by institutional planning, standardized selection criteria, and multidisciplinary follow-up.

### 4.6 Potential benefit to patients

The data suggest that the structured implementation of neuromodulation procedures could benefit a relevant portion of the population monitored in the service. With 893 patients eligible in four years, corresponding to an average of approximately 223 patients per year, there is a significant contingent of individuals with refractory motor complications, refractory tremor or drug intolerance that could be evaluated for an additional therapeutic option.

In addition to the potential clinical benefits, including improvement of motor symptoms, reduction of medications, and improvement of quality of life, the economic analysis suggests a possible favorable impact on long-term care costs. However, the confirmation of this benefit would depend on prospective studies with patients undergoing the procedure and systematic clinical-economic follow-up.

### 4.7 Limitations of the study

This study has some limitations. First, this is a descriptive cross-sectional study, with no prospective follow-up of patients undergoing neuromodulation procedures. Second, the data were collected in a single center, which may limit the generalizability of the results to other populations. Third, the cost-effectiveness analysis used data from the literature and local estimates for some cost components, not necessarily reflecting actual costs observed after the implementation of the procedure in the service.

Despite these limitations, this study characterizes a large population of patients with Parkinson’s disease followed up at a tertiary referral center, demonstrating a high frequency of motor complications, greater clinical complexity, and a significant proportion of individuals potentially eligible for neuromodulation. The findings suggest that the structured implementation of this therapeutic strategy, associated with standardized selection criteria and multidisciplinary follow-up, could expand the treatment options for patients with advanced PD and have a potentially favorable economic impact.

## Data Availability

The data supporting the findings of this study are not publicly available because they contain clinical patient information, but may be made available by the corresponding author upon reasonable request and institutional approval, where applicable.

## Acknowledgments

The authors would like to thank the São Lucas Hospital and the Neurology Service team for their institutional support and collaboration in organizing the clinical data used in this study.

## Notes

**Conflict of Interest:** The authors declare no conflict of interest.

**Use of Artificial Intelligence:** AI-assisted technologies were used only for language editing, translation, formatting support, and administrative preparation. All scientific content, clinical data, analyses, interpretations, and the final manuscript were reviewed and approved by the authors. No AI-assisted technology was used to generate or analyze study data.

### Competing Interest Statement

The authors have declared no competing interest.

### Author Declarations

The Research Ethics Committee of Hospital Sao Lucas, Campo Largo, Parana, Brazil, ethically approved this study (Opinion No. 2024-001). The requirement for informed consent was waived because the study was based on a retrospective review of medical records.

